# A Query Taxonomy Describes Performance of Patient-Level Retrieval from Electronic Health Record Data

**DOI:** 10.1101/19012294

**Authors:** Steven R. Chamberlin, Steven D. Bedrick, Aaron M. Cohen, Yanshan Wang, Andrew Wen, Sijia Liu, Hongfang Liu, William R. Hersh

## Abstract

Performance of systems used for patient cohort identification with electronic health record (EHR) data is not well-characterized. The objective of this research was to evaluate factors that might affect information retrieval (IR) methods and to investigate the interplay between commonly used IR approaches and the characteristics of the cohort definition structure.

We used an IR test collection containing 56 test patient cohort definitions, 100,000 patient records originating from an academic medical institution EHR data warehouse, and automated word-base query tasks, varying four parameters. Performance was measured using B-Pref. We then designed 59 taxonomy characteristics to classify the structure of the 56 topics. In addition, six topic complexity measures were derived from these characteristics for further evaluation using a beta regression simulation.

We did not find a strong association between the 59 taxonomy characteristics and patient retrieval performance, but we did find strong performance associations with the six topic complexity measures created from these characteristics, and interactions between these measures and the automated query parameter settings.

Some of the characteristics derived from a query taxonomy could lead to improved selection of approaches based on the structure of the topic of interest. Insights gained here will help guide future work to develop new methods for patient-level cohort discovery with EHR data.

## 1. Background

The intent of this research is to define and test a query taxonomy, applied to patient cohort definitions, which can explain the performance variations seen when retrieving these cohorts from electronic health record (EHR) data using automated methods. Also of interest is the possible relationship between a cohort taxonomy and different methods of retrieval and associated parameter settings. Patient cohort discovery in health records is an important task that is often used in academic institutions for research purposes [1]. This can be a very labor-intensive task requiring time spent to design custom queries for each cohort definition, or topic. Automated methods could improve the efficiency of this task, but have not been well-studied in this domain of information retrieval [2-5]. Promising research in medical record retrieval methods has been applied at the document or encounter level, used only structured data, used broad cohort definitions not applicable to research cohort identification, or generally focused on task definitions that differ from cohort identification, such as phenotyping [6-8]. For our purposes, cohort definitions not only contain disease diagnoses, but other complex features, such as lab tests, surgical procedures, medications, lab values, temporal relationships as well as combinations of structured and unstructured data.

Previous research has studied the performance of automated word-based queries used for complex patient-level cohort discovery with raw EHR data [5, 9], testing different parameter variations (n=48) for these queries against 56 complex patient cohort definitions, or topics. Performance was generally poor for these queries, with 86% of the topics having a median B-Pref [10] under .25 (scale 0-1) across the 48 query parameter variations. These queries also underperformed when compared to custom designed Boolean queries. There were also large performance variations between and within the 56 topics. The range of median B-Pref across topics was 0 at a minimum and .895 at a maximum, and within topics ranges were seen as small as .03 points and as large as .60 points. And finally, there were also differences in median B-Pref between the 48 query parameter settings, although these differences were not as dramatic as that seen from the for the topics.

The variation in performance seen across complex patient cohorts in previous research has led us to this research to explain, and predict, this variation by decomposing a patient-level cohort definition into a standard taxonomy. To do this we propose to use query performance data from previous research to test our taxonomy definitions. To our knowledge, this type of taxonomy has not been previously defined or tested for this type of cohort discovery task.

## 2. Materials and Methods

### 2.1 Test Data

To test the taxonomy developed for this project, we used the query performance data generated in a previous medical IR study [5]. This data was created using the Cranfield IR evaluation methodology [11] and trec_eval program [12] (**Fig 1)**. We used the B-Pref statistic as the performance measure for our evaluation. This statistic measures how many relevant patients were retrieved before non-relevant patients in ranked lists, and is also used when relevance judging is incomplete.

**Figure 1.**
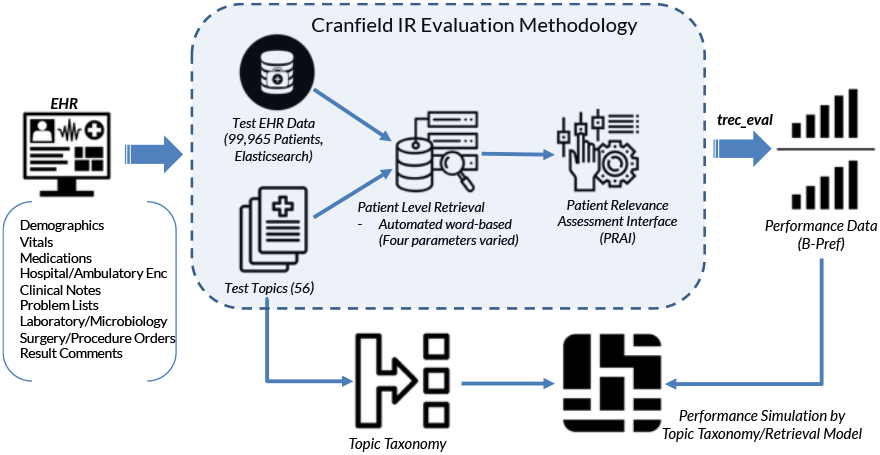
Overview of the generation of performance data used to test and model the query taxonomy definition.

Patient data originated from an Epic (Verona, WI) EHR system. A total of 99,965 unique patients with 6,273,137 associated encounters were stored in the Elasticsearch (v1.7.6) IR platform to evaluate the retrieval methods. There were a variety of document types associated with each patient: demographics, vitals, medications (administered, current, ordered), hospital and ambulatory encounters with associated attributes and diagnoses, clinical notes, problem lists, laboratory and microbiology results, surgery and procedure orders, and result comments. These patients had to have at least three primary care encounters between 2009 and 2013.

The 56 topics were derived from actual patient cohort requests seen at two major medical research institutions. Four examples of topic representations are seen in **Table 1**.

**Table 1.** Examples of four cohort definitions, selected from the total of 56, used to generate the query performance data.

|  |  |
| --- | --- |
| Adults with IBD who haven't had GI surgery | Adults with inflammatory bowel disease who haven't had surgery involving the small intestine, colon, rectum, or anus. |
| Adults with a Vitamin D lab result | Adults with a lab result for 25-hydroxy Vitamin D collected between May 15 and October 15. |
| Postherpetic neuralgia treated with topical and systemic medication | Adults with postherpetic neuralgia ever treated by concurrent use of topical and non-opioid systemic medications. |
| Children seen in ED with oral pain | Children who were seen in the emergency department with herpetic gingivostomatitis, herpangina or hand, foot, and mouth disease, tonsillitis, gingivitis, or ulceration (aphthae, stomatitis, or mucositis) not due to chemotherapy or radiation. |
| ACE inhibitor-induced cough | Adults who have used an ACE inhibitor and experienced ACE inhibitor-induced cough. |

Four parameters for the word-based queries were varied to create 48 total iterations (queries) for each of the 56 topics. The four parameters are as follows:

1. Topic Representation – <u>A</u> (summary statement), <u>B</u> (clinical case), or <u>C</u> (detailed criteria)
2. (Text Subset – only clinical <u>notes</u> or <u>all</u> document types (including structured data reporting as text)
3. Aggregation Method – patient relevance score calculated by summation (<u>sum</u>) of all documents or by maximum (<u>max</u>) value
4. Retrieval Model – BM25, also known as Okapi [13], Divergence from randomness (DFR) [14], Language modeling with Dirichlet smoothing (LMDir) [15], Default Lucene scoring, based on the term frequency-inverse document frequency (TF*IDF) model [16]

Random samples were selected from each run parameter iteration for each topic for manual relevance judgment by clinically trained reviewers. After the relevance assessment, final performance statistics were generated with the trec_eval program.

The final dataset contained the B-Pref performance statistic for all combinations of topics and query run parameters (56 topics x 48 run iterations = 2,688 unique queries).

### 2.2 Topic Taxonomy Characteristics

Our first step to explain and predict the performance of the word-based queries, was to create a topic taxonomy composed of 59 features. Three of the authors, who were trained clinically, iteratively developed a list of features that covered inclusion or exclusion of medical diagnoses and classifications, medications, procedures, lab tests, clinician information, patient demographics, information about the clinical setting, temporal measures and other aspects. Each of the 56 topics were then classified by these 59 features by the same three individuals. Fleiss Kappa was used to test interrater reliability [17].

We wanted to examine the association between the query performance, as measured by B-Pref, and the 59 taxonomy characteristic classifications of the 56 topics. To do this we did an exploratory data analysis by comparing one heatmap, clustered by query performance, to a second heatmap, clustered by characteristic assignment, with the topic clustering maintained from the first heatmap. The first heatmap used B-Pref as the statistic and clustered run parameter sets by topic. The second heatmap used the level of rater agreement (0-3) as the statistic and clustered by the taxonomy characteristic while maintaining the topic clusters found in the first performance-based heatmap. These heatmaps were compared for pattern similarities between performance clustering and taxonomy clustering.

### 2.3 Topic Taxonomy Structural Binary Features

To simplify the taxonomy definitions, focus more strictly on structural complexity, rather than content, representation, and to create features for model development and statistical testing, we created six binary features by grouping some of the 59 taxonomy characteristics into categories. At least one reviewer had to identify the taxonomy characteristics used to define these features as relevant to the topic. The determination of these components was based on our experience and knowledge of how these topic characteristics impact the complexity of query design. The overall goal was to investigate the relationship between these six taxonomy features and query performance, and to test for any performance related interactions between these features and the four word-based query parameters (topic representation, text subset, aggregation method, and retrieval model). These interactions capture the relationship between interventions (word-based query parameters) and inherent topic structure related to complexity (binary taxonomy features).

We used a beta regression model for this investigation. This model was trained on the B-Pref performance data with the four word-based query parameters, the six binary taxonomy features and all first-order interactions between the parameters and the features. Due to data limitations we felt that model coefficients and tests of significant might not be generalizable. We instead chose to use this model to predict B-Pref on all possible permutations of values of the parameters and features, and to investigate the patterns of the predicted B-Pref in this parameter/feature space. Since this simulated data contained all possible combinations of values of the four word-based parameters and the six binary taxonomy features, there were a total of 3,072 entries. Using the simulated data, we estimated the effect of the six binary taxonomy features individually, and the effect of the Topic Representations. We also used this simulated data for an exploratory data analysis, using a heatmap, to assess more complex interactions between the parameter space (interventions) and the binary feature space (inherent topic structure).

A beta regression mean model was selected because the response variable, B-Pref, is continuous, restricted to the unit interval [0,1], and asymmetrically distributed. The logit link function was used for these analyses. The regression was done with R (v3.3.1) using the package betareg (v3.1-2).

## 3. Results

### 3.1 Taxonomy Analysis – 59 Characteristics

**Table 2** lists the 59 characteristics of the query taxonomy, while **Table 3** lists the six binary features derived from the 59 taxonomy characteristics. We found moderate, substantial or almost perfect agreement by Fleiss kappa on 50 of the 56 topics, rated by the three clinically trained raters for the 59 query taxonomy characteristics (**Fig 2**).

**Table 2.** Taxonomy characteristics.

|  |  |
| --- | --- |
| Medical condition explicitly present? | Medical diagnosis (exc)? ICD or Text |
| Systems: CV | Medical diagnosis (inc)? ICD or Text |
| Systems: GI | Medication (inc)? |
| Systems: Neuro/Psych | Medication (exc)? |
| Systems: Dental | Labs? |
| Systems: Accident | Labs? (if yes) Values also? |
| Systems: Ophthalmic | Imaging? |
| Systems: Metabolic | Imaging? (if yes) Interpretation also? |
| Systems: Infectious | Physical exam? |
| Systems: GU | Physical exam? (if yes) Values also? |
| Systems: Pulmonary | Procedures/Surgeries (inc)? |
| Systems: Hematology | Procedures/Surgeries (exc)? |
| Systems: Reproductive | Location? (none) |
| Systems: Oral | Location? (inpatient) |
| Systems: Skin | Location? (outpatient) |
| Systems: Cancer | Location? (emergency) |
| Systems: Breast | Location? (ICU) |
| Systems: Immunology | Provider Type |
| System: Endocrine | Visit Type |
| System: Musculoskeletal | Visit Specialty |
| Age: none | Status? (deceased) |
| Age: adult | Status? (living) |
| Age: child | Temporal: sequence of events |
| Age: senior | Temporal: age at first diagnosis |
| Sex: none | Temporal: time with diagnosis |
| Sex: female | Temporal: time on medication |
| Sex: male | Temporal: date of procedure |
| Free text descriptions required, simple term? | Temporal: age at encounter |
| Free text descriptions required, complex concept? | Temporal: date of procedure/lab |
| Number of encounters |  |

**Table 3.**
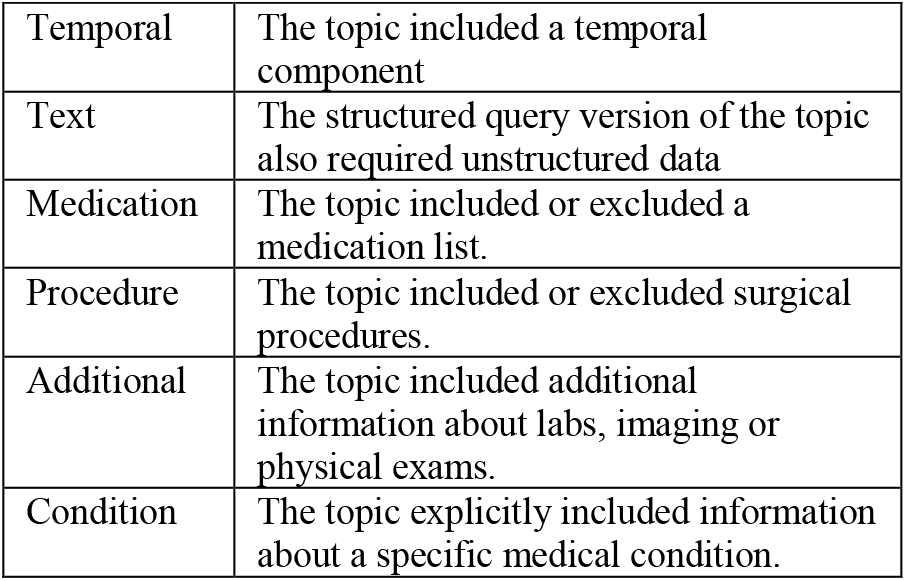
Taxonomy binary features.

**Figure 2.**
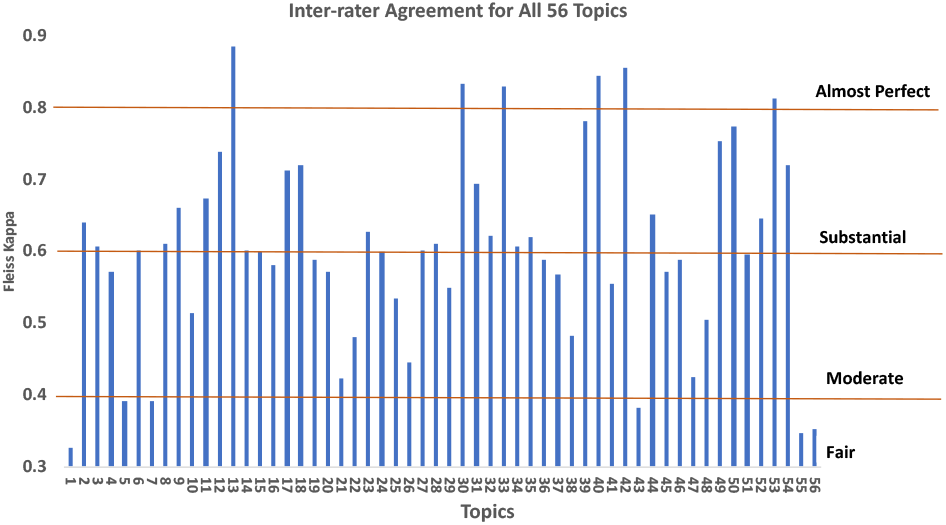
Interrater agreement for the 59 taxonomy characteristics applied to 56 topics. Fleiss Kappa was calculated for each topic based on agreement between three clinical trained raters on the 59 taxonomy characteristic assignments.

We next used heatmaps to investigate the relationship between word-based query performance and assignment to the 59 taxonomy characteristics (**Fig 3**). Topic clusters, based on B-Pref performance (left heatmap), were maintained for taxonomy characteristics (right heatmap), but column clustering was allowed for this heatmap. Performance-based clustering of topics can be seen for the B-Pref heatmap, but there do not appear to be similar patterns found in the taxonomy heatmap, while maintaining the same topic order. There does not appear to be an association between performance and the 59 taxonomy characteristics.

**Figure 3.**
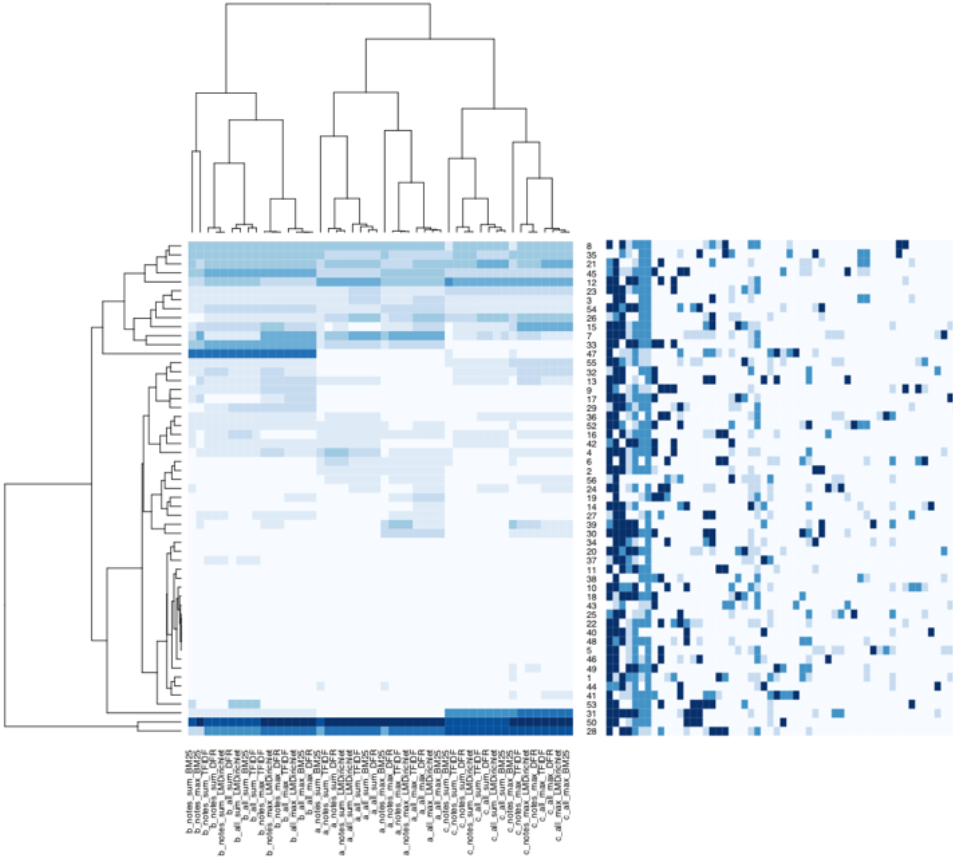
Comparison B-Pref Performance and Taxonomy Characteristic Clustering. The heatmap on the left are the topics by word-based query parameter settings clustered for B-Pref performance. Darker colors represent higher B-Pref values. The heatmap on the right maintains topic order from B-Pref clustering on the left (rows), but the columns, which represent the 59 taxonomy characteristic assignments, are clustered. Darker colors on the right heatmap represent the level of interrater agreement, but the lightest color means no association between the characteristic and the topic.

### 3.2 Taxonomy Analysis – 6 Binary Features

The first binary feature was positive if there was a temporal component in the topic (‘Temporal’, y/n). The 56 topics contain a variety of temporal conditions, including age at first diagnosis, time with diagnosis, chronological order of disease onset for several diagnoses, and medication use before or after first diagnosis. The second binary feature was positive if the topic could not be written exclusively with structured data, required some free text (‘Text’, y/n). The third binary feature was positive if the topic required a medication list check, either exclusions or inclusions or both (‘Medication’, y/n). The fourth binary feature was positive if there was a procedure in the topic. This includes any surgical or non-surgical procedure (‘Procedure’, y/n). The fifth binary feature was positive if additional values were required from lab tests, imaging, or physical exams. (‘Additional’, y/n). And finally, the sixth binary feature was positive if the topic required a specific disease diagnosis or diagnoses. Some topics were defined for cohorts who only received certain screening tests without an explicit disease requirement (‘Condition’, y/n). For our data, the beta regression model output did show that five of the six binary taxonomy features were associated with poorer performance, as measured by B-Pref. One feature, ‘text’, was associated with better performance. Features associated with poorer performance were designed to capture increased topic complexity, so this result is not surprising. The feature ‘text’ captures the ability of purely structured data to describe a medical topic, with or without added free text. Our result indicates that topics that require text, in addition to structured data, might perform better. And there were notable interactions between the taxonomy features and the run parameters, particularly between the feature ‘temporal’ and Topic Representation. Interestingly this analysis did not point to any notable interactions between the four word-based parameters. But it is not clear if these results are generalizable due to the specific nature of our 56 topic descriptions.

We then used the beta regression model, containing the four word-based parameters, six binary taxonomy features and the interactions between the parameters and features, to predict B-Pref with a simulated dataset. This dataset contained all possible permutations of the ten predictors. We varied each of the six binary taxonomy flags independently, while holding all other values constant, to estimate the impact of these flags. We also did this for Topic Representation (**Fig 4**). We again saw that five of the six binary taxonomy features were associated with poorer performance, and the feature ‘text’ was associated with improved performance. We also saw Topic Representation B associated with improved performance.

**Figure 4.**
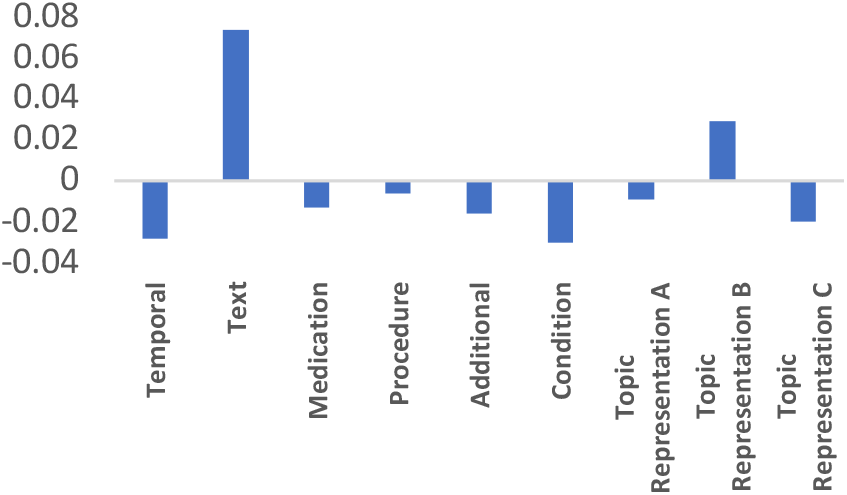
B-Pref prediction changes on simulated data for selected characteristics. Values represent change in predicted B-Pref using a simulated dataset with all permutations of the four word-based query parameters and the six binary taxonomy flags. All other values are held constant while varying the characteristic listed.

We also created a heatmap of the predicted B-Pref values generated from the simulated data (**Fig 5**). The x axis contained all possible permutations of the four word-based query parameters and the y axis contained all possible permutations of the six binary taxonomy features, and hierarchical clustering was done in both dimensions. Clear patterns of performance clustering can be seen, particularly around the combinations of three of the binary taxonomy features, temporal, text and condition. These three features are conceptually different from the other three (medication, procedure, additional) in that the latter are simple additions of information but the former represent more complex topic structural aspects. In addition, within specific combinations of these flags there are also clear variations in performance across different word-based parameter settings. In the bottom horizontal cluster, the best performance for topics without a temporal, text and condition component is seen with a completely different set of parameter settings than for topics with all three of these structural components. This performance pattern is an example of a possible interaction between the parameters and features, which could help guide the selection of parameters to optimize retrieval results.

**Figure 5.**
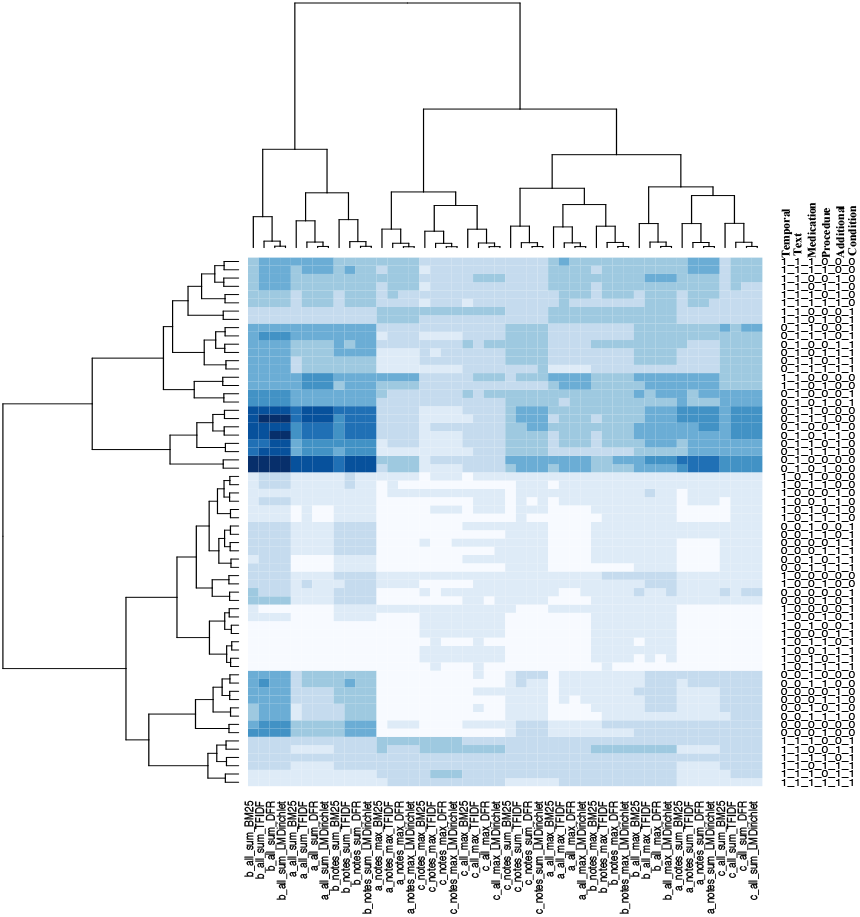
B-Pref predictions on simulated data. Darker values represent higher predicted B-Pref. Clustering was done on both axes. The x axis represents all permutations of the four word-based query parameters and the y axis represents all permutations of the six binary taxonomy features.

## 4. Discussion

The findings in our previous research, and the performance variation within and across topics, led us to pursue two further methods to understand and improve our results. In an attempt to understand our results, we developed a taxonomy for the topics that we hoped would identify characteristics associated with the differences in results. We first developed an exhaustive 59 parameter taxonomy that did not reveal any associations. However, when we reduced the taxonomy to six binary variables, we did find association with performance. As also shown by comparable work at Mayo Clinic [18], it may be possible with further prospective analysis that query taxonomy might lead to selection of different query approaches based on characteristics of the topic.

There is some evidence that applying a query taxonomy might improve performance. Further work with methods such as machine learning might yield improvements, although it is not clear what features will lead to performance improvement across varying topical criteria for different queries.

There were a number of limitations to this work. Our records were limited to a single academic medical center. There are many additional retrieval methods we could have assessed, but we would not have the resources to carry out the additional relevance judgments required as those additional methods would add new patients to be judged. It is also difficult to generalize our results due to the specificity of the topics. This will always be a limitation for this type of work since it would be extremely difficult to represent all possible cohort requests that could be seen for all forms of medical research. Finally, there is a global limitation to work with EHR data for these sorts of use cases in that raw, identifiable patient data is not easily sharable such that other researchers could compare their systems and algorithms with ours using our data.

## Data Availability

Our data contains patient health information and cannot be made publicly available.

## ACKNOWLEDGMENTS

This work was supported by NIH Grant 1R01LM011934.

